# Diagnostic accuracy and limit of detection of ten malaria parasite lactate dehydrogenase-based rapid tests for *Plasmodium knowlesi* and *P. falciparum*

**DOI:** 10.1101/2022.06.24.22276842

**Authors:** Angelica F. Tan, Sitti Saimah binti Sakam, Giri S. Rajahram, Timothy William, Mohammad Faruq Abd Rachman Isnadi, Sylvia Daim, Bridget E Barber, Steven Kho, Colin J. Sutherland, Nicholas M. Anstey, Seda Yerlikaya, Donelly A van Schalkwyk, Matthew J. Grigg

## Abstract

**Background:** *Plasmodium knowlesi* causes zoonotic malaria across Southeast Asia. First-line diagnostic microscopy cannot reliably differentiate *P. knowlesi* from other human malaria species. Rapid diagnostic tests (RDTs) designed for *P. falciparum* and *P. vivax* are used routinely in *P. knowlesi* co-endemic areas despite potential cross-reactivity for species-specific antibody targets.

**Methods:** Ten RDTs were evaluated: nine to detect clinical *P. knowlesi* infections from Malaysia, and nine assessing limit of detection (LoD) for *P. knowlesi (PkA1-H.1)* and *P. falciparum* (*Pf3D7*) cultures. Targets included *Plasmodium*-genus parasite lactate dehydrogenase (pan-pLDH) and *P. vivax* (*Pv*)-pLDH.

**Results:** Samples were collected prior to antimalarial treatment from 127 patients with microscopy-positive PCR-confirmed *P. knowlesi* mono-infections. Median parasitaemia was 788/µL (IQR 247-5,565/µL). Pan-pLDH sensitivities ranged from 50.6% (95% CI 39.6–61.5) (SD BIOLINE) to 87.0% (95% CI 75.1–94.6) (First Response^®^ and CareStart™ PAN) compared to reference PCR. *Pv*-pLDH RDTs detected *P. knowlesi* with up to 92.0% (95% CI 84.3-96.7%) sensitivity (Biocredit™). For parasite counts ≥200/µL, pan-pLDH (Standard Q) and *Pv*-pLDH RDTs exceeded 95% sensitivity. Specificity of RDTs against 26 PCR-confirmed negative controls was 100%. Sensitivity of the 6 highest performing RDTs were not significantly different when comparing samples taken before and after (median 3 hours) antimalarial treatment. Parasite ring stages were present in 30% of pre-treatment samples, with ring stage proportions (mean 1.9%) demonstrating inverse correlation with test positivity of Biocredit™ and two CareStart™ RDTs.

For cultured *P. knowlesi*, CareStart™ PAN demonstrated the lowest LoD at 25 parasites/µL; LoDs of other pan-pLDH ranged from 98 to >2000 parasites/µL. *Pv*-pLDH LoD for *P. knowlesi* was 49 parasites/µL. *P. falciparum*-pLDH or histidine-rich-protein-2 channels did not react with *P. knowlesi*.

**Conclusion:** Selected RDTs demonstrate sufficient performance for detection of all human malaria species including *P. knowlesi* in co-endemic areas where microscopy is not available, particularly for higher parasite counts, although cannot reliably differentiate among non-*falciparum* malaria.

## Introduction

The emergence of the simian malaria parasite *Plasmodium knowlesi*, known to infect humans across most of Southeast Asia, poses significant challenges to regional malaria control programs and World Health Organization (WHO) human malaria elimination progress (1). In Malaysia, a substantial increase in *P. knowlesi* cases has been reported since 2008 (2, 3), and *P. knowlesi* is now almost the sole cause of malaria with over 2600 documented cases in 2020 (4). Other countries with a higher burden of human malaria, such as Indonesia (5), India (6), Cambodia (7), Myanmar (8), and Thailand (9) have also reported *P. knowlesi* cases using molecular methods, with these cases initially being misidentified as other species using routine light microscopy. Due to similarities in morphology between *P. knowlesi* and other endemic *Plasmodium* species (10), there are major limitations to microscopy as the primary method of diagnosis in endemic areas, with regional prevalence likely underestimated (11, 12). Sensitive and specific malaria diagnostic tools are vital to ensure timely diagnosis, particularly in areas of high prevalence (13) where misidentification of *P. knowlesi* as other *Plasmodium* species can lead to delayed or inappropriate treatment and increased risk of fatal outcome (14).

The utilization of antibody panels for human malaria has enabled the design of antigen-capture rapid diagnostic tests (RDTs) as an alternative to microscopy (15). Although no RDT has been designed for specific detection of *P. knowlesi*, the current list of WHO-prequalified *in vitro* diagnostics (IVD) for malaria includes seven tests that can detect the genus-conserved parasite lactate dehydrogenase (pLDH) antigen (16). However, the performance of these RDTs for detecting *P. knowlesi* infections has not been investigated. Moreover, in the last round of the WHO-FIND Malaria RDT Evaluation Programme published in 2018, composite test positivity was considerably increased compared to earlier rounds, with the best-performing pLDH-based tests for *P. falciparum* and *P. vivax* detection approaching 100% even at low parasite counts of 200/µL (17). McCutchan *et. al* previously detailed a panel of monoclonal antibodies targeting multiple epitopes against LDH isoforms of the major human *Plasmodium* species in addition to *P. knowlesi*. Three epitopes with nominal specific binding capacity to LDH from *P. vivax* and *P. falciparum* were found to cross-react with *P. knowlesi* LDH, although not with LDH from other simian malaria species (18). However, previous studies evaluating the utility of a small number of *P. vivax* and *P. falciparum*-pLDH based RDTs from previous WHO RDT testing rounds demonstrated poor sensitivity for the detection of clinical *P. knowlesi* infections (19–21).

Within Southeast Asia, there is large variance in the use of microscopy or RDTs as the first-line malaria diagnostic tool, in addition to the specific type of RDTs targeting different antigens such as pLDH or *P. falciparum*-specific histidine-rich protein 2 (HRP2) commercially sourced between endemic countries. For example, Malaysia uses microscopy as the first-line detection tool, whereas a variety of different malaria RDTs are primarily used within certain areas of other countries such as Indonesia (1). At a regional level, India is the biggest consumer of RDTs as part of their national malaria control strategy due to the prevalence of both *P. falciparum* and *P. vivax* (22, 23). Although parasitological diagnosis using RDTs is recommended by the WHO where microscopy is unavailable, many countries still face logistical and cost difficulties in providing access to RDTs in remote areas, compounded by the limited analytical sensitivity of pLDH-based RDTs, with cases potentially treated or referred to tertiary centers based on clinical suspicion (24–28).

In this study we evaluate the utility of commercially-available RDTs for detecting clinical *P. knowlesi* infections from an endemic setting. We also estimate the limit of detection of RDTs using dilution series of *P. knowlesi* and *P. falciparum* laboratory cultures (29).

## Methods

### Rapid diagnostic tests selection

The study included all seven WHO-prequalified malaria RDTs that target pan-pLDH (16). Based on the WHO round 8 malaria RDT product testing results available at the time of the study design (17), two additional RDTs were selected according to the best panel detection score for both *P. falciparum* and *P. vivax* at parasite count thresholds of 200 and 2000 parasites/µL. RDTs with a lower false positivity rate were then preferentially included. Subject to source availability from manufacturer, the final ten RDTs were selected (**Table 1**). Eight RDTs were evaluated on both clinical *P. knowlesi* samples and laboratory cultured *P. knowlesi* and *P. falciparum* isolates. One RDT (careUS™) was tested only on clinical samples, and another (Meriscreen OneStep™) on only laboratory samples.

**Table 1.**
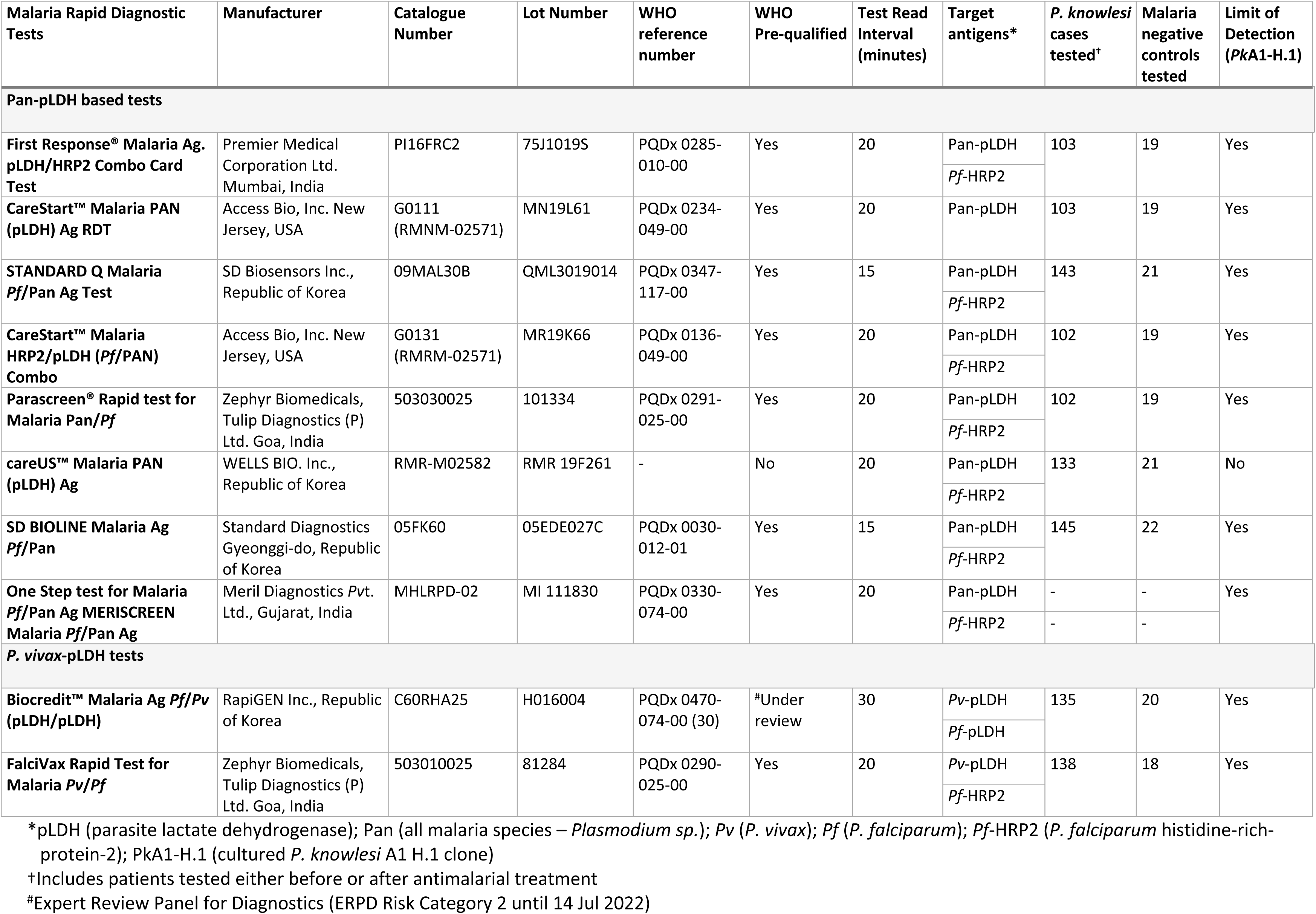
Product information, target antigens and clinical data for the ten rapid diagnostic tests evaluated for *P. knowlesi* detection

### Study sites, subjects, and ethical approval

Patients were enrolled from Ranau District Hospital in Sabah, Malaysia, a secondary referral center servicing the administrative district (area of 3609 km^2^), accepting referrals from 16 primary health clinics, including all cases of malaria for in-patient management. Patient blood samples were collected and demographic and clinical data were recorded on standardized case record forms as part of an ongoing prospective observational malaria study. Patients presenting to the hospital study site were included if they were positive for *P. knowlesi* by microscopy, were more than 1 year of age, and provided appropriate written informed consent. Malaria-negative individuals presenting with or without a fever at the same healthcare facility were also prospectively enrolled as controls. The study was approved by the Medical Research and Ethics Committee of the Ministry of Health, Malaysia (NMRR-10-584-6684) and Menzies School of Health Research, Australia (HREC 10-1431).

### Blood sample procedures

Venous whole blood samples were collected in EDTA vacutainers prior to antimalarial treatment where possible. All patients had thick and thin blood films prepared from pre-treatment whole blood for verification of parasitaemia by microscopy. Microscopic quantification of *P. knowlesi* parasitaemia (parasites per microlitre) was conducted by an experienced research microscopist equivalent to WHO Level 1 competency; parasitaemia was calculated from the number of parasites per 200 white blood cells on thick blood film, multiplied by the individual patient’s total white cell count (31) obtained from routine hospital laboratory flow cytometry. Specific blood volumes specified in manufacturers’ instructions were used to evaluate the RDTs. The remaining EDTA whole blood samples were stored at −80°C for subsequent malaria species confirmation by reference real time PCR.

### *Plasmodium* species confirmation by reference real-time PCR

All microscopically diagnosed malaria cases and microscopy-negative controls were tested by PCR for *P. knowlesi, P. falciparum, P. vivax, P. malariae* and *P. ovale* spp. In brief, genomic DNA was extracted from 200 μL of whole blood using QIAamp DNA Blood Mini Kits (Cat. No.: 51106; QIAGEN) according to the manufacturer’s manual, with a final elution volume of 200 μL. PCR detection of *Plasmodium* species was performed by laboratory research members blinded to the microscopy results. A real-time PCR assay, QuantiFast™ targeting the 18S SSU rRNA gene was conducted once for each sample in accordance with the manufacturers’ and Sabah Public Health Laboratory protocols, using the Bio-Rad CFX96 Touch™ PCR machine (Bio-Rad, USA). QuantiFast™ real-time PCR was carried out over two separate reactions, with the first reaction for *Plasmodium* genus screening, followed by subsequent *P. knowlesi* and other human *Plasmodium* species-specific detection if positive (32).

### RDTs conducted on clinical *P. knowlesi* samples

The nine available antigen-based RDTs were transported and stored with appropriate conditions and temperature monitoring. All RDTs were conducted according to the manufacturers’ instructions at Ranau District Hospital in Sabah. Test results were independently interpreted by two readers, with a positive result documented when both the result and control test strip lines were deemed to be present. In the event of a result discrepancy, a third individual blinded to the initial results performed a tie-break reading. The intensity of each test band on the RDT was graded visually on a scale from 0 to 4 according to a standard color chart provided by FIND, with 0 indicating no line observed and 4 indicating the highest intensity line observed. A threshold score of ≥ 1 was used to indicate a confirmed parasite signal. Due to staggered enrolments and differences in timing of procurement, RDTs were not all obtained simultaneously and tested on exact sample sets. Key clinical and demographic parameters were tested across samples used for individual RDTs to ensure no confounding group differences.

### Limit of detection (LoD) for *in vitro P. knowlesi* and *P. falciparum*

Cultures of *P. knowlesi* (PkA1-H.1) and *P. falciparum* (*Pf*3D7) parasites were maintained *in vitro* as described previously (33). Parasite dilutions for RDT testing were performed in whole blood obtained from either the UK National Health Service Blood and Transplant (NHSBT) or Cambridge Bioscience. Negative controls (whole blood without *Plasmodium* parasites) and positive controls (whole blood containing *P. falciparum* parasites at 2,000 parasites/µL) were supplied by FIND, the global alliance for diagnostics. For RDT testing, *P. knowlesi* or *P. falciparum* cultured parasites were diluted two-fold in whole blood from 2,000 parasites/µL. All controls and test samples were applied on RDT cassettes according to the manufacturer’s instructions. Experiments were performed in duplicate at the London School of Hygiene & Tropical Medicine, UK, and repeated on four separate occasions for *P. knowlesi* and between two to four times for *P. falciparum*. Signals were assessed and classified visually according to the same RDT color chart supplied by FIND for the clinical evaluation. The threshold used to indicate a confirmed parasite signal present was a consistent faint line with a score of ≥ 1. The careUS™ pan-pLDH RDT was not included in the LoD evaluation due to procurement difficulties from the manufacturer at the time of testing.

### Statistical analysis

The primary analysis compared the diagnostic accuracy of each RDT to detect microscopy-positive, PCR-confirmed *P. knowlesi* clinical infections collected prior to antimalarial treatment. Diagnostic accuracy was evaluated using the number of true positive, false negative, false positive and true negative results to calculate sensitivity and specificity (34). Exact binomial confidence intervals (CI) of 95% for each diagnostic metric were calculated and reported. Diagnostic positive and negative predictive values incorporated the estimated prevalence of malaria, calculated from the proportion of febrile patients with a positive microscopy result presenting to the district hospital site during the study period (35). Independent comparisons between the sensitivity of RDT test components were conducted using Fisher’s exact test for: a) parasitaemia of above or below 200 parasites/µL, and b) samples collected before versus after administration of antimalarial treatment. For the latter, logistic regression was used to further compare the sensitivity of RDT antigen targets when controlling for median time in hours post treatment. Group differences across RDTs compared log normalized age of participants and parasite count distribution using one-way ANOVA, and sex using Chi-square test. Interobserver agreement between RDT line intensity readers was measured using the kappa coefficient (36). Spearman’s correlation test was used to assess the association between line intensity gradings and parasitaemia as well as correlation between proportion of early trophozoite (ring) stages to RDT test positivity. The limit of detection (LoD) measurements with 95% confidence intervals were estimated using logistic regression analysis in R (version 4.0.5, the R Foundation for Statistical Computing, 2020). All other statistical analyses were carried out using STATA v16 (TX, USA).

## Results

### Baseline demographics

From July 2020 to August 2021, 1,698 febrile patients presented to the hospital site and were screened by microscopy, with 508 identified as *P. knowlesi* infections. Of these, 492 were subsequently confirmed on PCR, resulting in a background malaria prevalence of 28.9% (95% CI 20.5-24.8%) in this cohort. A total of 202 microscopically determined *P. knowlesi* clinical cases and 26 malaria-negative controls were prospectively enrolled for RDT testing.

Of the *P. knowlesi*-positive samples with RDTs tested, 127 (63%) were collected from patients prior to anti-malarial treatment and were included in the primary analysis. The median age of patients with *P. knowlesi* malaria was 34 years (range 4 – 87 years). PCR-confirmed negative controls had a median age of 34 years (range 12 – 78 years) and included 21 (81%) febrile patients. Most malaria patients and controls were male (79% and 69%, respectively). There were no statistically significant differences in age or sex between malaria cases and negative controls across the RDTs conducted.

The geometric mean parasite count of *P. knowlesi* infected patients was 734 parasites/µL (95% CI 563 – 956), with a range of 18 – 41,883 parasites/µL. Of 202 *P. knowlesi* enrolled patients, 55 (27%) had a parasite count of less than 200/µL, which included 26 (47%) with samples collected prior to treatment. Three patients were classified with severe *P. knowlesi* malaria using WHO research criteria (37). There were no statistically significant differences in the distribution of parasite counts used for individual RDT testing.

### Sensitivity of pan-pLDH based RDTs for P. knowlesi detection

The target pan-pLDH antigen made up the major component of seven RDTs tested using clinical isolates (**Table 1**). The First Response^®^ and CareStart™ PAN RDTs contained the highest performing pan-pLDH tests to detect clinical *P. knowlesi* infections, with a sensitivity of 87.0% (95% CI 75.1 - 94.6) and lowest detected parasitaemia of 50/µL for both (**Table 2**). The poorest performing pan-pLDH based RDT was the SD Bioline with a sensitivity of 50.6% (95% CI 39.6 - 61.5), and a parasitemia threshold for positivity of 254 parasites/µL. The median parasitaemia of patients with a false-negative pan-pLDH result was 260 parasites/µL (IQR 125 – 737). False-negative results were reported at high parasite counts for the SD Bioline (8,315 parasites/µL) and careUS™ (6,341 parasites/µL) RDTs.

**Table 2.**
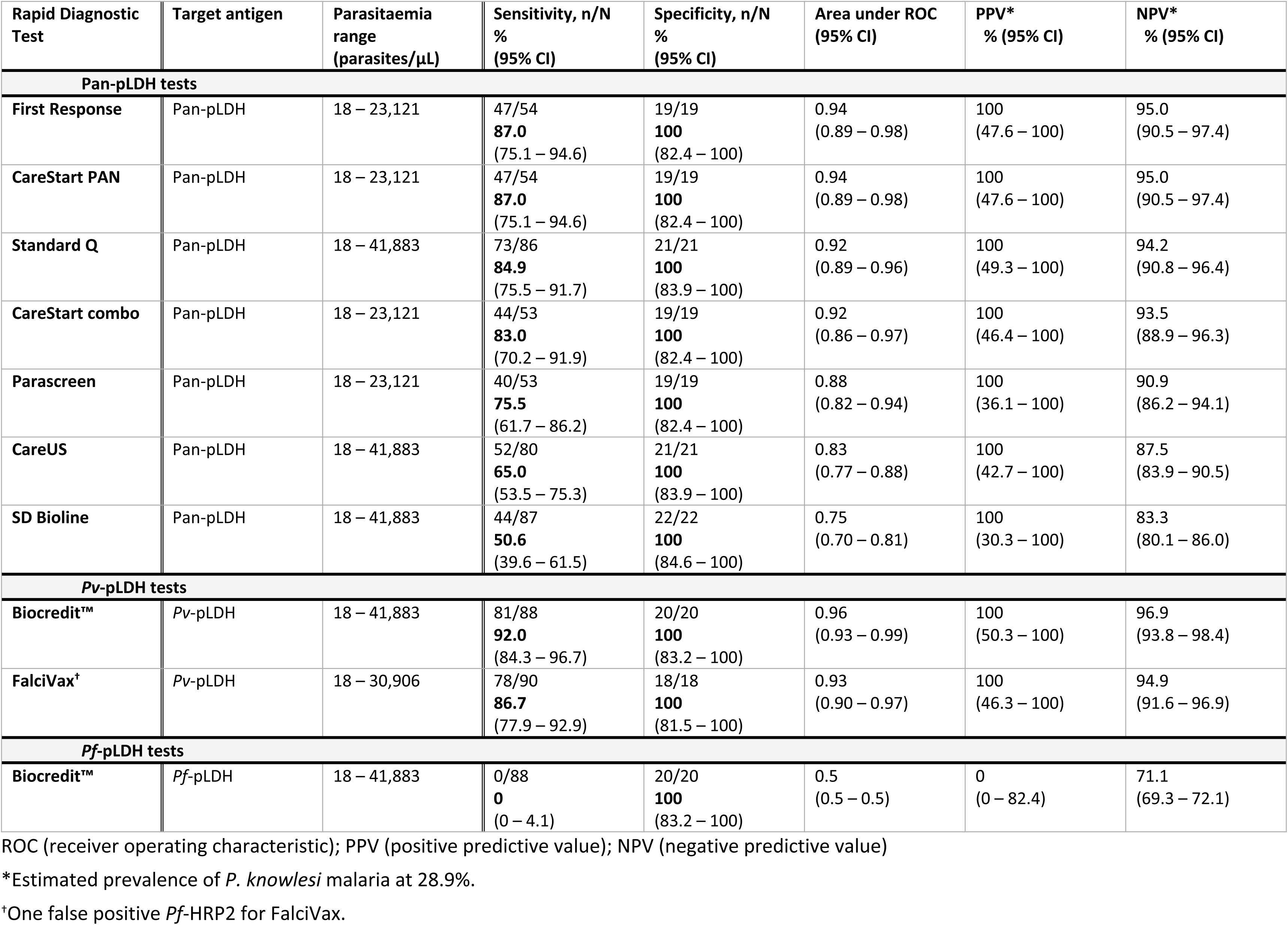
Performance of RDTs in detecting *P. knowlesi* clinical infections (prior to antimalarial treatment)

| Rapid Diagnostic Test | Target antigen | Parasitaemia range (parasites/μL) | Sensitivity, n/N % (95% CI) | Specificity, n/N % (95% CI) | Area under ROC (95% CI) | PPV* % (95% CI) | NPV* % (95% CI) |
| --- | --- | --- | --- | --- | --- | --- | --- |
| **Pan-pLDH tests** |  |  |  |  |  |  |  |
| **First Response** | Pan-pLDH | 18 – 23,121 | 47/54<br>**87.0**<br>(75.1 – 94.6) | 19/19<br>**100**<br>(82.4 – 100) | 0.94<br>(0.89 – 0.98) | 100<br>(47.6 – 100) | 95.0<br>(90.5 – 97.4) |
| **CareStart PAN** | Pan-pLDH | 18 – 23,121 | 47/54<br>**87.0**<br>(75.1 – 94.6) | 19/19<br>**100**<br>(82.4 – 100) | 0.94<br>(0.89 – 0.98) | 100<br>(47.6 – 100) | 95.0<br>(90.5 – 97.4) |
| **Standard Q** | Pan-pLDH | 18 – 41,883 | 73/86<br>**84.9**<br>(75.5 – 91.7) | 21/21<br>**100**<br>(83.9 – 100) | 0.92<br>(0.89 – 0.96) | 100<br>(49.3 – 100) | 94.2<br>(90.8 – 96.4) |
| **CareStart combo** | Pan-pLDH | 18 – 23,121 | 44/53<br>**83.0**<br>(70.2 – 91.9) | 19/19<br>**100**<br>(82.4 – 100) | 0.92<br>(0.86 – 0.97) | 100<br>(46.4 – 100) | 93.5<br>(88.9 – 96.3) |
| **Parascreen** | Pan-pLDH | 18 – 23,121 | 40/53<br>**75.5**<br>(61.7 – 86.2) | 19/19<br>**100**<br>(82.4 – 100) | 0.88<br>(0.82 – 0.94) | 100<br>(36.1 – 100) | 90.9<br>(86.2 – 94.1) |
| **CareUS** | Pan-pLDH | 18 – 41,883 | 52/80<br>**65.0**<br>(53.5 – 75.3) | 21/21<br>**100**<br>(83.9 – 100) | 0.83<br>(0.77 – 0.88) | 100<br>(42.7 – 100) | 87.5<br>(83.9 – 90.5) |
| **SD Bioline** | Pan-pLDH | 18 – 41,883 | 44/87<br>**50.6**<br>(39.6 – 61.5) | 22/22<br>**100**<br>(84.6 – 100) | 0.75<br>(0.70 – 0.81) | 100<br>(30.3 – 100) | 83.3<br>(80.1 – 86.0) |
| **Pv-pLDH tests** |  |  |  |  |  |  |  |
| **Biocredit™** | Pv-pLDH | 18 – 41,883 | 81/88<br>**92.0**<br>(84.3 – 96.7) | 20/20<br>**100**<br>(83.2 – 100) | 0.96<br>(0.93 – 0.99) | 100<br>(50.3 – 100) | 96.9<br>(93.8 – 98.4) |
| **FalciVax†** | Pv-pLDH | 18 – 30,906 | 78/90<br>**86.7**<br>(77.9 – 92.9) | 18/18<br>**100**<br>(81.5 – 100) | 0.93<br>(0.90 – 0.97) | 100<br>(46.3 – 100) | 94.9<br>(91.6 – 96.9) |
| **Pf-pLDH tests** |  |  |  |  |  |  |  |
| **Biocredit™** | Pf-pLDH | 18 – 41,883 | 0/88<br>**0**<br>(0 – 4.1) | 20/20<br>**100**<br>(83.2 – 100) | 0.5<br>(0.5 – 0.5) | 0<br>(0 – 82.4) | 71.1<br>(69.3 – 72.1) |

ROC (receiver operating characteristic); PPV (positive predictive value); NPV (negative predictive value)
\*Estimated prevalence of *P. knowlesi* malaria at 28.9%.247 †One false positive *Pf*-HRP2 for FalciVax.

### Cross-reactivity and sensitivity of P. vivax-pLDH to detect P. knowlesi

The *Pv*-pLDH target antigen of the Biocredit™ RDT demonstrated a sensitivity of 92.0% (95% CI 84.3 - 96.7), the highest for detecting *P. knowlesi* among all RDTs tested. The Biocredit™ RDT also had the lowest recorded parasite counts of 32 parasites/µL and 40 parasites/µL for clinical samples tested either before or after antimalarial treatment, respectively (**Table 3**). The *Pv*-pLDH test component of the FalciVax RDT also cross-reacted with *P. knowlesi*, albeit with a slightly lower sensitivity of 86.7% (95% CI 77.9 - 92.9) and lowest detected clinical parasitaemia of 50 parasites/µL. The median parasitaemia of patients with a false-negative *Pv*-pLDH result was 83 parasites/µL (IQR 41 – 203), with the highest false-negative result for Biocredit™ recorded at 251 parasites/µL and for FalciVax at 422 parasites/µL.

**Table 3.** Limit of detection (LoD) for nine rapid diagnostic tests.

| Rapid Diagnostic Test | In vitro Limit of Detection<br>Parasites/μL [95% CI] |  |  | Lowest recorded positive result<br>Parasites/μL |
| --- | --- | --- | --- | --- |
|  | *PkA1-H.1* culture isolate*<sup>#</sup> | *Pf3D7* culture isolate<sup>‡</sup> |  | *P. knowlesi* clinical sample<sup>°</sup> |
|  | Pan-pLDH | Pan-pLDH | HRP2 | Pan-pLDH |
| **First Response** | 98.03<br>[89.66 - 106.40] | 98.09<br>[89.68-106.49] | <7.8<br>[n/a] | 50 |
| **CareStart™ PAN** | 24.63<br>[18.20 - 31.05] | 49.18<br>[41.75-56.62] | n/a | 50 |
| **STANDARD Q** | 195.61<br>[186.26-204.97] | 778.73<br>[767.39-790.09] | 24.67<br>[18.21-31.13] | 40 |
| **CareStart™ combo** | 98.03<br>[89.66 - 106.40] | 195.69<br>[186.29-205.08] | 12.39<br>[6.90-17.87] | 50 |
| **Parascreen®** | 195.61<br>[186.26 - 204.97] | 195.69<br>[186.29-205.08] | <7.8<br>[n/a] | 50 |
| **One Step test MERISCREEN** | 1555.14<br>[1543.24 - 1567.05] | 390.48<br>[380.08-400.89] | 24.67<br>[18.21-31.13] | - |
| **SD BIOLINE** | >2000<br>[n/a] | 1556.54<br>[1544.88-1568.19] | 12.39<br>[6.90-17.87] | 254 |
|  | *Pv*-pLDH | *Pf*-pLDH | *Pf*-HRP2 | *Pv*-pLDH |
| **Biocredit™** | 49.18<br>[41.75 - 56.62] | 49.18<br>[41.75-56.62] | n/a | 40 |
| **FalciVax** | 49.18<br>[41.75 - 56.62] | n/a | 12.39<br>[6.90-17.87] | 50 |

\*No *P. falciparum*-HRP2 bands were detected with the *Pk A1-H.1* culture isolate for any of the RDTs screened (i.e., no cross-reactivity)
<sup>#</sup>No *P. falciparum* bands were detected with the *Pk A1-H.1* culture isolate for either of the RDTs screened, but cross reactivity was observed with *P. vivax* lines.
<sup>‡</sup>No *P. vivax* lines were detected with the *Pf3D7* culture isolate (i.e., no cross-reactivity)
<sup>°</sup>Includes patients tested before antimalarial treatment only

### Cross-reactivity of P. falciparum-specific antibodies with P. knowlesi

Amongst the RDTs tested, only the Biocredit™ RDT included a *Pf*-pLDH component for which no cross-reactivity was observed for any of the *P. knowlesi* clinical samples tested. Of the seven RDTs which contain a *Pf*-HRP2 component, a single false-positive result was recorded for the FalciVax RDT at a *P. knowlesi* count of 31 parasites/µL (Table 2). Six other false positive *Pf*-HRP2 results were observed when the following RDTs were tested on samples collected after antimalarial treatment: Parascreen^®^ (150/µL), First Response (148/µL and 1,924/µL) and FalciVax (49/µL, 162/µL and 608/µL).

### Sensitivity of RDT detection at different levels of P. knowlesi parasitaemia

Of *P. knowlesi*-infected patients whose samples were collected prior to treatment, 26 (20%) had a parasite count less than 200/µL, with a median parasitaemia of 88 parasites/µL (IQR 50 – 133) (**Supplementary Table 1**). Consistent with the overall data, the best performing test in detecting samples with low parasitaemia was *Pv*-pLDH from the Biocredit™ RDT with a sensitivity of 73.7% (95% CI 48.8 to 90.9%), followed by the FalciVax *Pv*-pLDH test at 50%. The pan-pLDH tests performed poorly at these low parasite counts, with the SD Bioline unable to detect a single *P. knowlesi*-positive sample and both the CareStart™ PAN and First Response^®^ tests demonstrating a low sensitivity of 42.9% (95% CI 9.9 - 81.6%); **Figure 1**.

**Figure 1.**
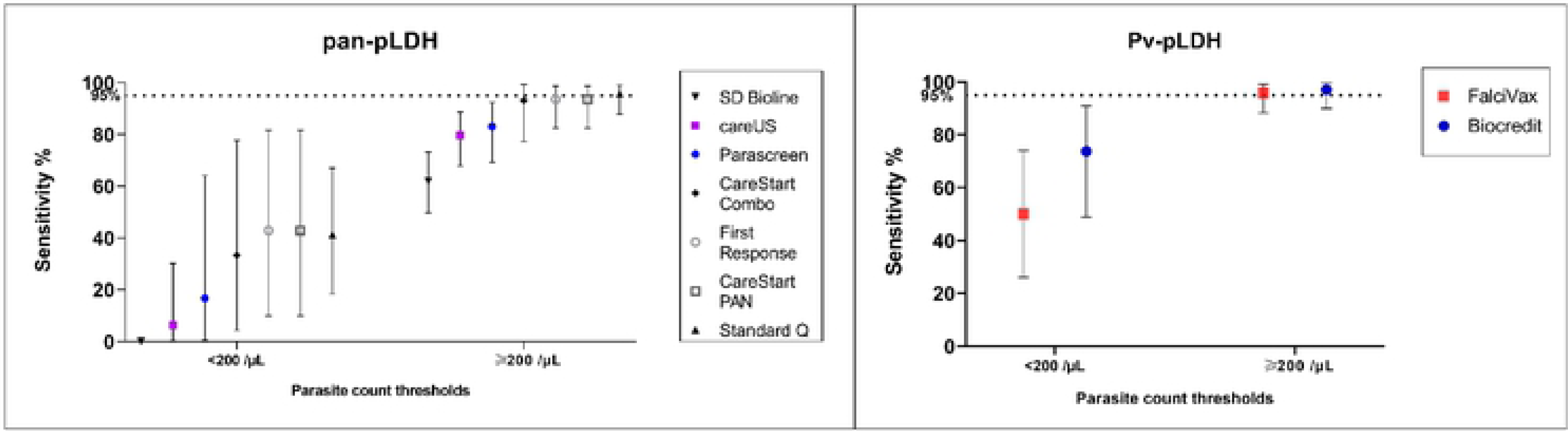
Sensitivity (and 95% confidence intervals) of (left) pan-pLDH and (right) Pv-LDH in detecting P. knowiesi for parasite count groups of <200/μL and ≥200/μL

The parasite count group of 200 parasites/µL and above comprised of 101 (80%) *P. knowlesi*-infected participants, with a median parasitaemia of 1,245/µL (IQR 489 – 6,224). Both pan-pLDH and *Pv*-pLDH targets demonstrated excellent utility for *P. knowlesi* detection, with three RDTs exceeding the predefined sensitivity threshold of 95% (38) at which performance is considered sufficient for clinical use (Figure 1). These RDTs included the *Pv*-pLDH component of the Biocredit™ and FalciVax RDTs with sensitivities of 97.1% (95% CI 89.9 - 99.6%) and 95.8% (95% CI 88.3 – 99.1%) respectively, in addition to the pan-pLDH of Standard Q at 95.7% sensitivity (95% CI 87.8 – 99.1%). Comparable performance was seen for the pan-pLDH of the CareStart™ PAN and First Response^®^ RDTs with a sensitivity of 93.6% for both. Performance further improved for the 74 (58.3%) *P. knowlesi* malaria cases with parasitaemia of more than 500 parasites/µL, with the same five highest performing RDTs all recording 100% sensitivity. The SD Bioline pan-pLDH test also performed poorest in this parasite count group with a sensitivity of 62.0%.

### Proportion of ring stages affect test positivity

Microscopic evaluation of parasite life-stages were carried out on the 127 clinical samples with *P. knowlesi* infections collected prior to antimalarial treatment. The mean proportions of early trophozoite (rings), late trophozoite and schizont asexual life-stages in a single infection were 1.9% (95% CI 0.8 – 2.9), 92.8% (95% CI 90.0 – 95.6) and 5.3% (95% CI 2.7 – 8.0) respectively. Asynchronous life-stages were found in 52% (66/127) of *P. knowlesi* infections, with synchronous late trophozoites and schizonts present in 46% and 2% respectively. The proportion of ring stages inversely correlate with test positivity for CareStart™ combo, CareStart™ PAN and Biocredit™ RDTs (Spearman’s rho < −0.32; p<0.002); **Supplementary Table 2**. Logistic regression controlling for parasitaemia demonstrated that the probability of a pan-pLDH or *Pv*-pLDH positive test outcome decreases by 0.9% (95% CI 0.8 – 1.0) for CareStart™ combo (p=0.044) and 0.8% (95% CI 0.7 – 1.0) for Biocredit™ (p=0.016) with each percentage increase in mean ring stage proportion. In comparison, *in vitro* cultures were all asynchronous with geometric mean proportions of rings and late trophozoites of 59.1% (95%CI 49.7 - 70.4) and 37.6% (95%CI 28.8 - 49.0) respectively.

### Specificity against malaria-negative samples

All pan-pLDH and *Pv*-pLDH RDT tests exhibited 100% specificity for malaria diagnosis, as demonstrated by the lack of response for all malaria-negative controls in individually tested RDTs. Two false-positives were recorded for *Pf*-HRP2 antibody, one each by First Response^®^ and Parascreen^®^ respectively (**Table 2**).

### Performance of RDTs to detect sub-microscopic P. knowlesi infections

Five patients excluded from the primary analysis were initially enrolled as microscopy-negative febrile controls; however, subsequent reference PCR detected *P. knowlesi*. Sample DNA was re-extracted and PCR repeated for confirmation, and research microscopy cross-checked 500 slide fields to confirm the infection was below both routine and research standard microscopic limit of detection. A single submicroscopic *P. knowlesi* infection was positive on a pan-pLDH test (CareStart™ PAN) and the *Pv*-pLDH based RDTs (FalciVax and Biocredit™). The other 4 submicroscopic *P. knowlesi* infections were negative on all RDTs. Inclusion of these submicroscopic *P. knowlesi* infections in the primary analysis would not have resulted in statistically significant differences in reported RDT sensitivities.

### RDT detection for P. knowlesi in samples collected before versus after anti-malarial treatment

RDTs were evaluated independently on 75 *P. knowlesi*-infected patients after administration of weight-based antimalarial treatment with oral artemether-lumefantrine, including 6.5% of whom initially received at least 1 dose of intravenous artesunate. RDTs were conducted a median of 3.3 hours (IQR 1.7 - 7.1 hours) after antimalarial treatment, with no statistically significant difference in time among the evaluated RDTs. Of the seven RDTs with pan-pLDH as the target antigen, two had significantly higher sensitivities when tested on pre-versus post-treatment samples: SD Bioline (50.6% vs 29.3% respectively; p = 0.01) and Parascreen^®^ (75.5% vs 55.1%; p = 0.03) (**Figure 2)**. Diagnostic sensitivity for all primary target antigens when categorized to parasite count groups of below and above 200/µL was not otherwise affected by whether samples were taken before or after administration of anti-malarial treatment (**Supplementary Table 3**).

**Figure 2.**
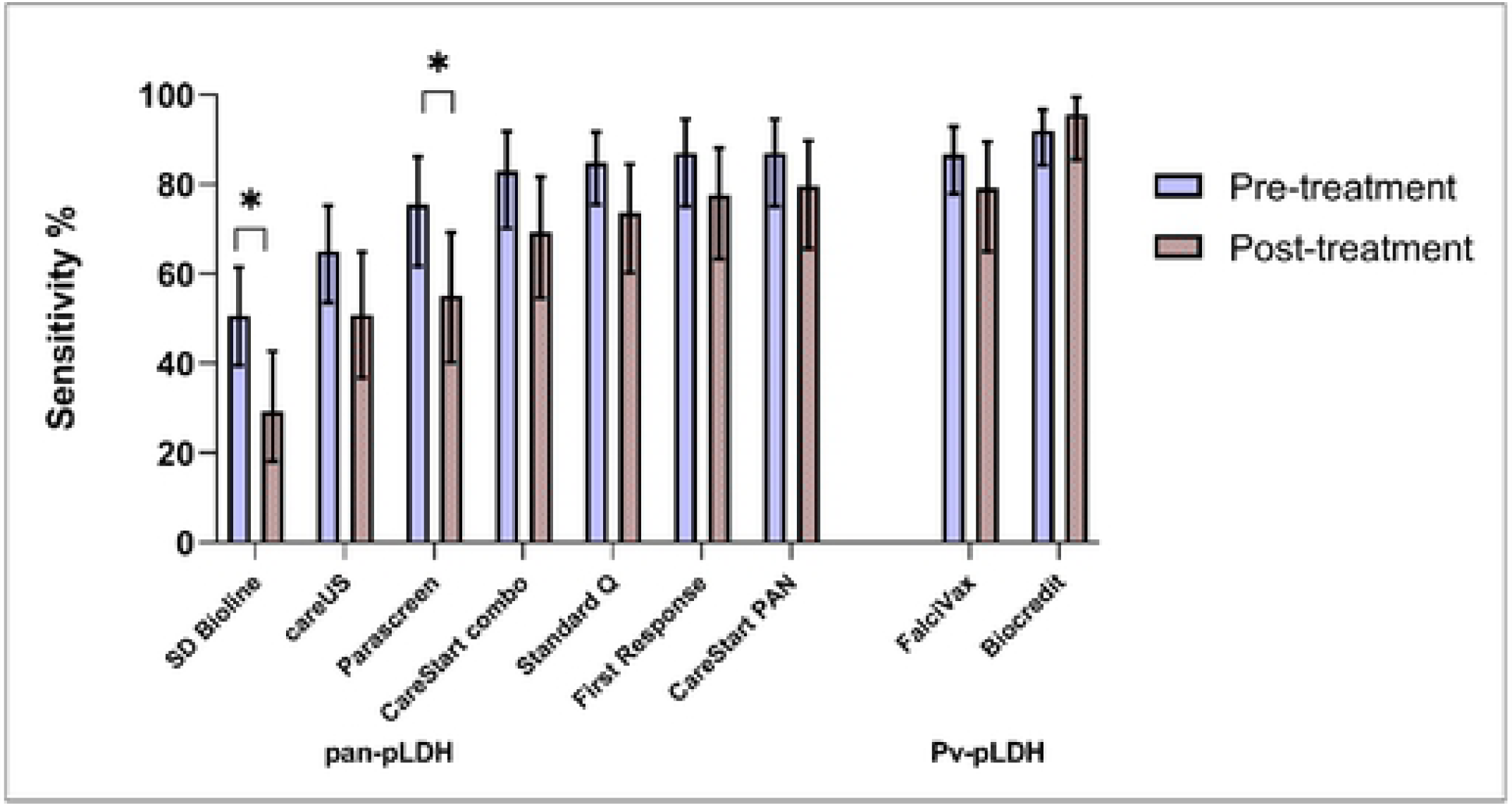
Comparative sensitivity of RDT targets (pre versus post antimalarial treatment) for P. knowlesi including 95% confidence intervals (*) indicate p<0.05.

### RDT line intensity and interobserver agreement

Line intensity gradings positively correlated with increasing *P. knowlesi* parasitaemia, including for both pan-pLDH and *Pv*-pLDH target antigens (Spearman’s rho >0.73; p<0.001) (Figure 3). Reporting of binary results for the primary pan-pLDH and *Pv*-pLDH antigens for all nine RDTs demonstrated excellent interobserver agreement with kappa values of >0.90. For each individual RDT, overall interobserver agreement was assessed for line intensity grades 1-4, indicative of independently reproducible readings for each colour intensity. Pan-pLDH-based tests recorded kappa values ranging from minimal (κ=0.40; SD Bioline) to moderate (κ=0.77; First Response^®^) agreement, whereas the *Pv*-pLDH lines gave weak kappa values of 0.50 (Biocredit™) and 0.57 (FalciVax).

**Figure 3.**
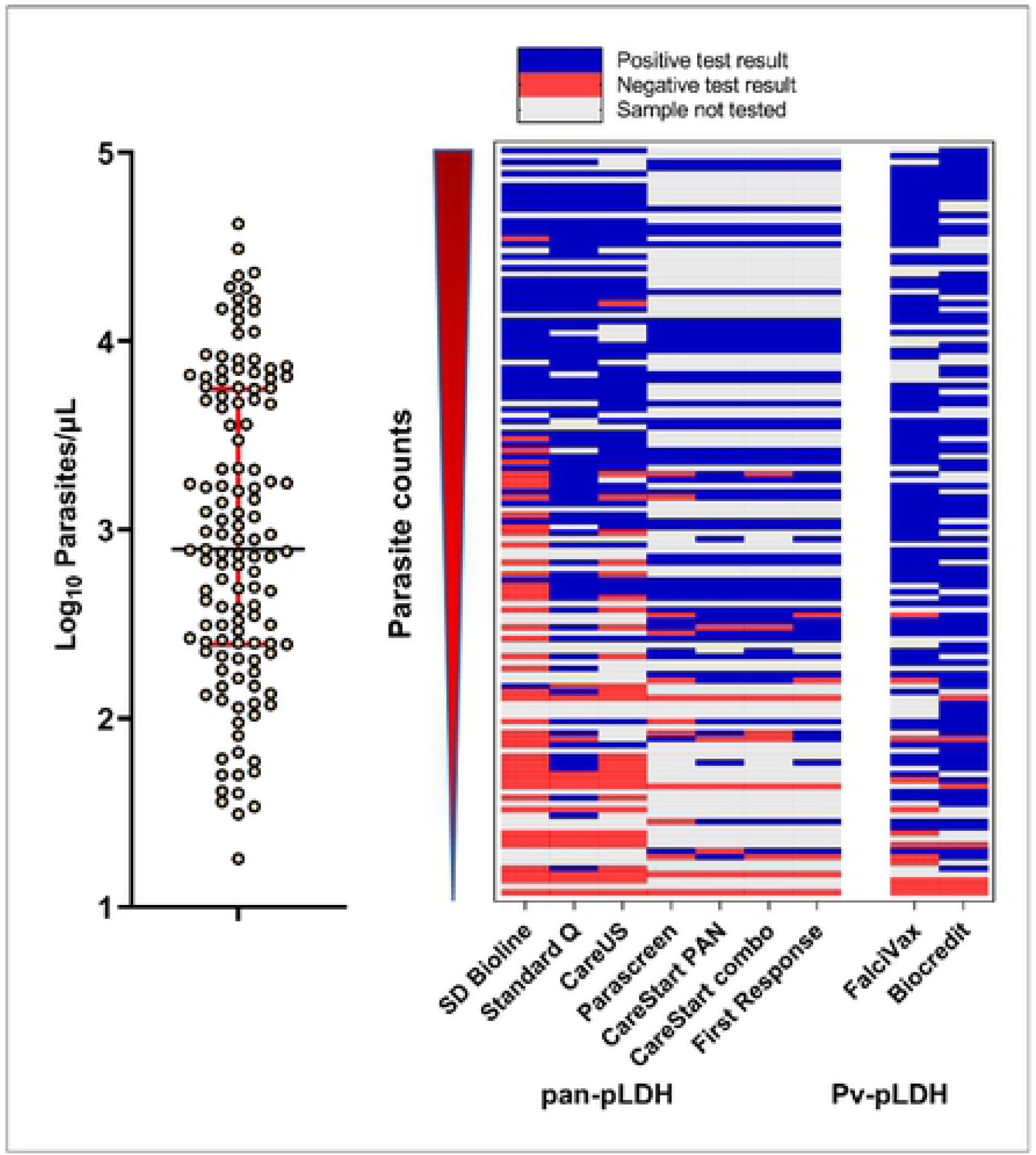
(Left) Distribution of parasite counts of clinical samples tested. (Right) Overview of tested samples with increasing parasite counts and corresponding outcomes in the nine RDTs tested for detection of clinical *P knowlesi*.

### Detection limits for in vitro P. knowlesi and P. falciparum cultures

*P. knowlesi* and *P. falciparum* parasites cultured *in vitro* were detected by all pan-pLDH channels of the RDTs tested. For the detection of *PkA1-H.1* via the pan-pLDH channel, the most sensitive RDT was the CareStart™ PAN with a LoD of 25 parasites/µL, followed by CareStart™ combo and First Response^®^ with a LoD of 98 parasites/µL for both (**Table 3**). The least sensitive RDTs were the Meriscreen One Step Test (LoD of 1,555 parasites/µL) and SD Bioline, with the latter not able to detect *PkA1-H.1* even at the highest parasite concentration tested of 2,000 parasites/µL. Similar to the *PkA1-H.1* dilutions, the pan-pLDH channel of CareStart™ PAN recorded the highest sensitivity for the *Pf3D7* cultures (LoD of 49 parasites/µL), whereas SD Bioline was the least sensitive (LoD of 1,556 parasites/µL). All RDTs with HRP2 channels were able to detect *Pf3D7* with LoD values of >25 parasites/µL.

Culture-derived *PkA1-H.1* were also detected by the *Pv*-pLDH channels of Biocredit™ and FalciVax^®^. No cross-reactivity occurred in the corresponding *P. falciparum*-pLDH or HRP2 channels, similar to test results for clinical samples. The LoD of cultured parasites in the *Pv*-pLDH channels were determined to be 49 parasites/µL for both RDTs, also consistent with the lowest parasitaemia detected from clinical samples (Table 3). In comparison, culture-derived *Pf3D7* was detected by *P. falciparum* channels of both the Biocredit™ (LoD 49 parasites/µL) and FalciVax (LoD 12 parasites/µL) RDTs, with no cross-reactivity occurring in the *Pv*-pLDH channels.

## Discussion

Rapid parasitological confirmation of malaria is a key pillar of effective clinical management recommended by the WHO in all endemic settings (39). Extending this rationale to include zoonotic malaria requires the availability of accurate point-of-care detection methods for *P. knowlesi*. Given the lack of routine microscopy available in many areas of Southeast Asia, the need to develop and validate RDTs for *P. knowlesi* diagnosis was identified as a key research priority in 2017 by the *P. knowlesi* WHO Evidence Review Group (40). The most sensitive antibody for *P. knowlesi* detection in this study was the anti-*Pv*-pLDH component of the Biocredit™ RDT at 92%, much higher than that seen in previously published RDT evaluations encompassing earlier iterations of *Pv*-pLDH antibodies (19–21). Among the RDTs with pan-pLDH as the primary antibody target, CareStart™ PAN and First Response^®^ were the most sensitive for detecting *P. knowlesi* clinical samples at 87%. No RDTs met the overall 95% sensitivity threshold deemed sufficient by the WHO to replace microscopy as the preferred first-line diagnostic tool (41). However, performance was excellent for parasite counts of more than 200/μL, with both *Pv*-pLDH based RDTs (Biocredit™ and Falcivax^®^) and one pan-pLDH based RDT (Standard Q) demonstrating sensitivity above 95%, increasing further to 100% for parasite counts more than 500/μL. These RDTs may have reasonable utility in areas where microscopy is not routinely available, although in endemic regions such as Sabah, Malaysia, symptomatic *P. knowlesi* infections commonly occur at low parasite densities, with previous large studies reporting around 12% of patients having a parasite count less than 200/μL (42).

The performance of WHO pre-qualified/Round 8 pre-validated RDTs used for detection of *P. knowlesi* in this study were markedly improved from previously evaluated RDTs selected from earlier WHO testing rounds, using similar pLDH targets with commercially undisclosed epitopes. Previous studies aiming to detect low-level *P. knowlesi* clinical monoinfections in Sabah using pan-pLDH (OptiMAL-IT) and VOM (non-*P. falciparum*)-pLDH (CareStart™) RDTs demonstrated poor sensitivities of less than 42% (19). Other pan-pLDH (First Response^®^, [Cat. No:II6FRC30]) and pan-*Plasmodium* aldolase (ParaHIT) RDTs also demonstrated insufficient sensitivities of 74% and 23%, respectively (20). A separate study in Sarawak, Malaysia, reported comparably low sensitivity values: 71% for pan-pLDH (OptiMAL-IT), 40% for pan-pLDH (Paramax-3) and 29% for aldolase (BinaxNOW^®^) (21). Test results with pLDH-based assays more broadly, including for *P. falciparum* detection, have been shown to have variable performance (41, 43) and are influenced by several factors including binding affinity of the target protein, parasite density in blood samples and the prozone effect (44–46). This reflects differing characteristics of the antibodies used by each manufacturer in these proprietary test platforms.

This study confirms the cross-reactivity of *P. knowlesi* LDH antigens with *Pv*-pLDH antibodies used in two commercially available RDTs tested (Biocredit™ and FalciVax^®^). *P. knowlesi* LDH has previously been shown to have high sequence identity (>96%) with the *P. vivax* ortholog (47), however the separate LDH isoforms exhibit both shared and unique surface residues associated with differential epitope binding of targeted antibodies (18). With both RDTs demonstrating high test positivity for *P. knowlesi* despite not being developed or previously validated for this purpose, specificity for *P. vivax* detection in co-endemic areas is likely to be significantly lower than manufacturers report. In areas where both species are endemic, misidentification could potentially lead to inappropriate clinical management, such as chloroquine for unsuspected severe knowlesi malaria (48), or unnecessary liver-stage treatment with primaquine for presumed *P. vivax* (49). However, in areas where *P. vivax* has been successfully eliminated, the cross-reactivity seen in *Pv*-LDH channels of both these RDTs could be used to accurately detect *P. knowlesi* infections and initiate appropriate antimalarial treatment.

Exploiting differences in the degree of cross-reactivity with pan-pLDH and *P. vivax* or *P. falciparum*-pLDH has also been hypothesized as a potential strategy to distinguish separate *Plasmodium* species. A recent study assessed quantified *Pv*-LDH cross-reactivity against both recombinant pLDH proteins and laboratory cultured isolates from *P. knowlesi*, in addition to the related simian species *P. cynomolgi* (50). *P. knowlesi*-pLDH recombinant protein cross-reacted with both the pan-pLDH and *Pv*-pLDH antibodies; however, the *P. cynomolgi*-pLDH protein only reacted with the latter. Genetically, *P. cynomolgi* is also closely related to *P. vivax* and likely shares high sequence identity with orthologous *Pv*-pLDH targets. In the current study, *P. knowlesi* parasites from both clinical and laboratory-cultured samples were also detected by all pan-pLDH and *Pv*-pLDH components tested, meaning they are likely unable to differentiate *P. vivax*. The quantified ratio of pan-to *Pv*-pLDH responses could provide additional information if this approach was used during the development of the RDTs (51), however this requires further investigation. With the exception of CareStart™ PAN, *in vitro* LoD values from the pan-pLDH based RDTs were generally higher than the lowest detected parasitaemia of the clinical samples. The presumed larger pLDH-to-parasite ratio in clinical whole blood samples could be occurring due to either higher proportions of late parasite stages with greater pLDH production compared to cultured samples or hypothesized elevated pLDH secreted into the peripheral circulation by hidden extravascular parasites, as seen in the spleen in *P. vivax* (52, 53). LDH expression levels may also differ between parasite species and needs to be considered as a potential source of variability in inter-species sensitivity, in addition to primary sequence variation.

The single *Pf*-pLDH based test (Biocredit™) evaluated in this study did not demonstrate any cross-reactivity with *P. knowlesi*, despite a *Pf*-pLDH containing RDT in a previous study demonstrating low sensitivities of up to 29% (19). This is consistent with previous modelling of pLDH isoforms which also demonstrated both shared and unique residues between *P. falciparum* and *P. knowlesi* associated with specific epitope antibody binding or lack thereof (18). For public health surveillance purposes including measuring progress towards elimination of *P. falciparum*, RDTs used in co-endemic areas would ideally need to differentiate between *P. falciparum* and *P. knowlesi*, given similarities in the microscopic morphology of their early ring-stages (10, 12). Correct identification of *P. falciparum* also allows appropriate transmission-blocking administration of single-dose primaquine, not required for *P. knowlesi* due to the gametocidal effect of current blood-stage treatments in this species (54, 55). In this context, the *Pf*-HRP2 antigen remains the preferred test component to differentiate non-falciparum malaria in areas such as Malaysia where hrp2/3 gene deletions have not been reported (19). When compared to previous studies of diagnostic accuracy for *P. knowlesi* which also reported false-positive results for *Pf*-HRP2 channels (19, 21), the proportion seen for *P. falciparum* was lower in the RDTs evaluated in this study. Additionally, no bands were observed for any RDT in the *Pf*-HRP2 channel with the cultured *PkA1-H.1*, further confirming the high specificity of this test for *P. falciparum* parasites.

The RDTs evaluated in this study showed variable performance for detection of *P. knowlesi*, with many demonstrating sufficient sensitivity for diagnostic use despite the inherent limitations in specificity against other *Plasmodium* species. Current efforts to develop novel malaria RDTs or modify those currently available to improve specificity for *P. knowlesi* and other clinically relevant simian malaria species face considerable obstacles (26) as manufacturers of existing commercial malaria RDTs do not often disclose which pLDH epitopes are bound by their proprietary antibodies (11). Current malaria RDT development is appropriately focused on developing highly-sensitive tests in order to facilitate elimination of *P. falciparum* and *P. vivax*, due to the increasingly recognized role that low-level asymptomatic infections have on sustaining transmission (56). However, simian malaria species such as *P. knowlesi* and *P. cynomolgi* are not routinely included as part of the RDT development or validation process. The inclusion of reliable *P. knowlesi-*specific antigen targets in RDTs with satisfactory sensitivity would play an important role in malaria case management, allow accurate assessment of progress towards WHO elimination goals for *P. falciparum* and *P. vivax*, and provide a vital epidemiological tool for understanding regional zoonotic malaria transmission and the true burden of disease, especially in resource-constrained areas.

## Conclusion

The findings of this study support utilising RDTs for initial rapid detection, but not identification, of knowlesi malaria in areas of higher endemism where access to microscopy is limited or unavailable. Improvement in the performance of pLDH-based RDTs for *P. knowlesi* detection was apparent with high sensitivities recorded particularly for parasite counts above 200/µL. When used as a single parasitological diagnostic test, malaria RDTs should incorporate a *Pf*-HRP2 component for non-falciparum species differentiation. The development of newer generation malaria RDTs to be deployed in *P. knowlesi* endemic areas, including through the WHO RDT product testing process, should include validation against *P. knowlesi* and other zoonotic *Plasmodium* species capable of human infection.

## Data Availability

All relevant data are within the manuscript and its Supporting Information files.

## Acknowledgements

We thank the study participants, the IDSKKS malaria research team (Maizatul Farina binti Abd Mutalib and Noorazela binti Mohamed Yassin), Ranau District Hospital director and clinical/laboratory staff, Dr. Fiona Binzini of Ranau Public Health District (PKD Ranau), Dr. Dyang Ruksuna Md Ruhul, Dr. Wan Mohamad Haziman Bin Wan Taib and Dr. Mohd Fakhrurizal Bin Matdiris as well as the Sabah State Public Health Laboratory. We would like to thank the Director General of Health Malaysia for the permission to publish this article.

This work was supported by the Australian Centre of Research Excellence in Malaria Elimination, the National Health and Medical Research Council, Australia (Grant Numbers 1037304 and 1045156, fellowships to NMA [1042072], MJG [1138860]), the National Institutes of Health, USA (R01AI160457-01) and Malaysian Ministry of Health (grant number BP00500/117/1002) awarded to GSR. AFT is supported via a Malaysia Australia Colombo Plan Commemoration (MACC) and Australian Government Research Training Program (RTP) Scholarship at Charles Darwin University. MJG is supported by the Australian Centre for International Agricultural Research, Australian Government (LS-2019-116). The analytical performance analysis part of this work was conducted with funds from the Bill & Melinda Gates Foundation (grant no. OPP1172683).

